# Genome-wide association and multi-omic analyses reveal new mechanisms for Heart Failure

**DOI:** 10.1101/19006510

**Authors:** Marios Arvanitis, Yanxiao Zhang, Wei Wang, Adam Auton, 23andMe Research Team, Ali Keramati, Neil C. Chi, Bing Ren, Wendy S. Post, Alexis Battle

## Abstract

Heart failure is a major medical and economic burden in the healthcare system affecting over 23 million people worldwide. Although recent pedigree studies estimate heart failure heritability around 26%, genome-wide association studies (GWAS) have had limited success in explaining disease pathogenesis. We conducted the largest meta-analysis of heart failure GWAS to-date and replicated our findings in a comparable sized cohort to identify one known and two novel variants associated with heart failure. Leveraging heart failure sub-phenotyping and fine-mapping, we reveal a putative causal variant found in a cardiac muscle specific regulatory region that binds to the ACTN2 cardiac sarcolemmal gene and affects left ventricular adverse remodeling and clinical heart failure in response to different initial cardiac muscle insults. Via genetic correlation, we show evidence of broadly shared heritability between heart failure and multiple musculoskeletal traits. Our findings extend our understanding of biological mechanisms underlying heart failure.

## Introduction

Heart failure is a highly prevalent disease [1] that constitutes a major medical and economic burden in the healthcare system, accounting for approximately 1-2% of the annual healthcare budget in developed countries [2]. Although almost any disease that directly or indirectly affects myocardial function can lead to the eventual development of clinical heart failure, it is well established that certain intrinsic homeostatic mechanisms like the renin-angiotensin-aldosterone axis and the sympathetic nervous system potentiate the effects of a variety of myocardial insults and cause adverse left ventricular remodeling [3], suggesting that multiple cellular mechanisms that lead to the disease are shared regardless of the inciting condition.

The increasing appreciation of an underlying strong heritable component of clinical heart failure further strengthens the argument for shared, yet unidentified, disease mechanisms whose discovery could reveal novel targets for its treatment and prevention. Indeed, large recent pedigree studies estimate heart failure heritability to be 26-34% [4]. However, large scale genome wide associations studies (GWAS) for heart failure have been unsuccessful to-date at uncovering a significant proportion of this estimated heritability underscoring a major unmet need in cardiovascular genetics. In fact, the largest published GWAS for heart failure to-date has only identified one genome-wide significant locus for all-comers with the disease that the investigators attribute to its overlap with atrial fibrillation [5], thereby revealing no actionable targets that predispose to heart failure development.

## Results and Discussion

### GWAS meta-analysis and replication identify novel loci associated with heart failure

We performed a large scale GWAS meta-analysis of five cohorts that study cardiovascular disease and two population genetics cohorts, all of European ancestry comprising a total of 10,976 heart failure cases and 437,573 controls. We used the 1000 Genomes phase 3 reference panel to impute variants from SNP array data and analyzed a total of 13,066,955 unique genotyped or high confidence imputed variants (INFO score > 0.7) with a minor allele frequency > 1%. We analyzed each individual cohort using a logistic mixed model and meta-analyzed all studies with fixed effects inverse-variance meta-analysis.

The combined meta-analysis revealed one previously identified and two novel loci associated with clinical heart failure at a genome-wide significance threshold (p-value < 5e-8) (Figure 1A, Table 1, Supplemental Table 1). All identified leading variants are common (MAF > 10%) and are located in non-coding regions of the genome (Supplemental Figure 1). We validated our genome wide significant loci in an independent cohort of 24,829 self-reported heart failure cases and 1,614,513 controls of European ancestry from the personal genetics company 23andMe, Inc. with all three sentinel variant associations successfully replicating at a nominal p-value level (p < 0.05) (Table 1) and after Bonferroni adjustment.

**Table 1.**
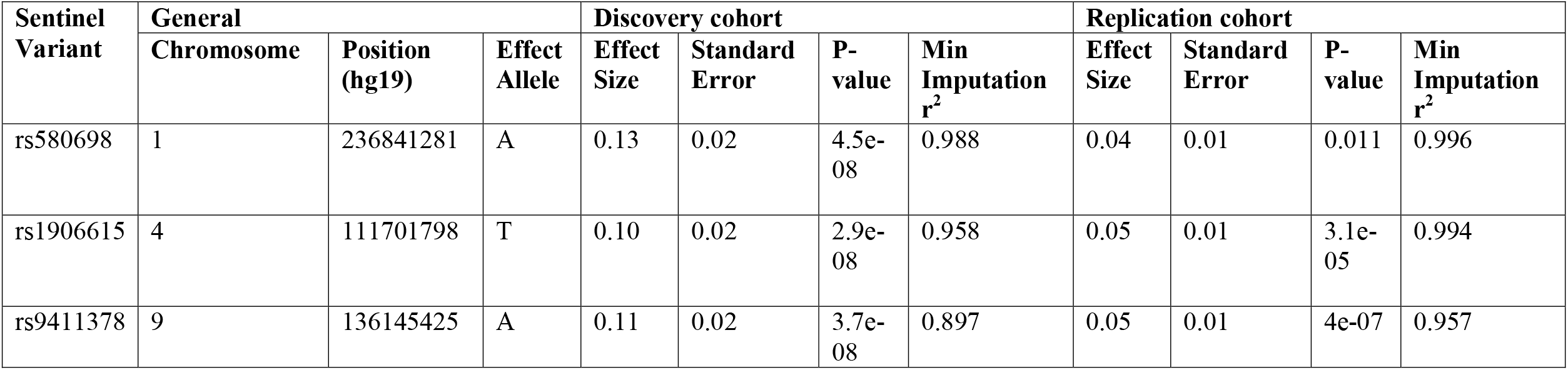
Lead variant associations in the Discovery and Replication cohorts

**Figure 1.**
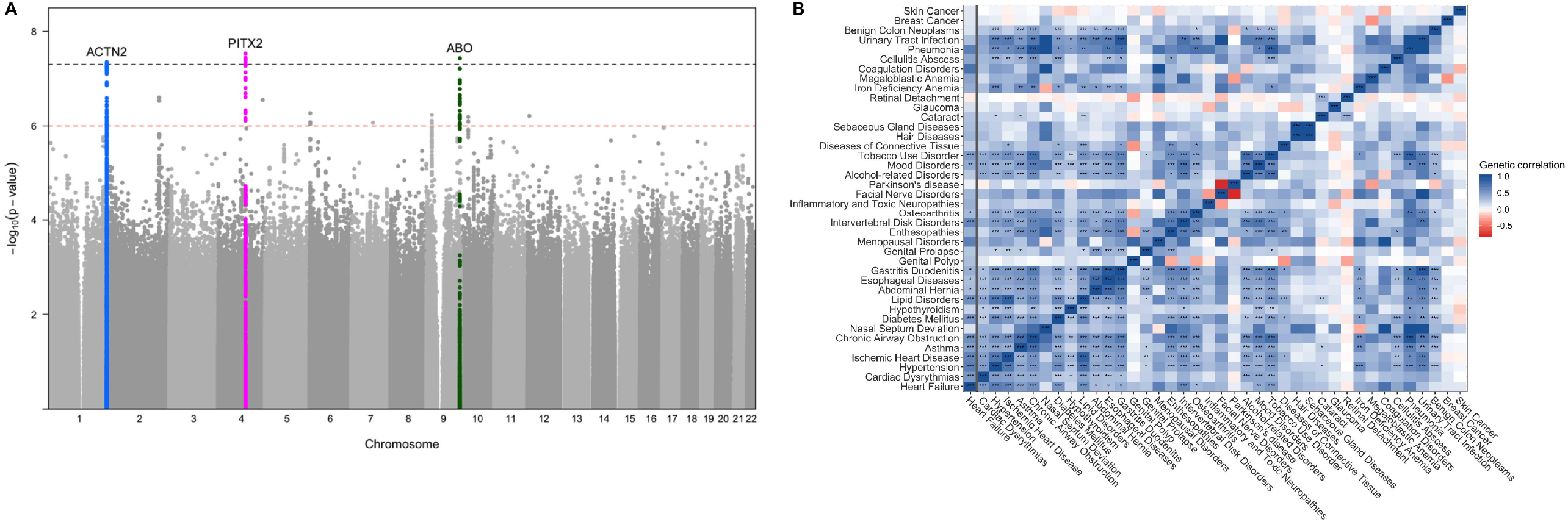
A. Manhattan plot of the GWAS meta-analysis. The black dotted line denotes the genome-wide significance threshold (p-value < 5e-8) while the red dotted line denotes the suggestive threshold (p-value < 1e-6). B. Genetic correlation values between Heart failure and other complex traits. The x-axis shows the genetic correlation r-value. Traits are color coded based on the group in which they belong. Stars denote the Bonferroni adjusted p-value: (*) 0.001 < p-value < 0.01, (**) 0.0001 < p-value < 0.001, (***) p-value < 0.0001

### Genetic correlation reveals shared heritability between heart failure and musculoskeletal traits

We subsequently performed linkage disequilibrium (LD) score regression to estimate heart failure heritability driven by common variants and the genetic correlation between heart failure and other complex traits. Liability scale SNP heritability for the disease assuming a population prevalence of 1.8% [6] was 5.9% (SE 0.7%), much lower than the pedigree-based estimates of 26%, a discrepancy that has been observed for other complex traits [7] and could be explained by multiple factors including rare variants. The LD score regression intercept was 0.99 indicating no inflation beyond what can be accounted for by polygenicity. As expected, we saw significant genetic correlation between heart failure and known heart failure risk factors such as hypertension, ischemic heart disease, adverse lipid profiles, diabetes, and atrial fibrillation. We also found a strong association between heart failure and pulmonary, musculoskeletal, and GI traits (Figure 1B, Supplemental Table 2).

### Atrial Fibrillation has a causal role in heart failure development

We investigated each genome-wide significant locus in depth. The chromosome 4 locus tagged by the SNP rs1906615 is found in an intergenic region close to the PITX2 gene. This locus was previously identified as containing the strongest evidence for association with atrial fibrillation [8] and has been reported as a significant locus in a recent heart failure GWAS [5]. However, that association was thought to be mediated via the relative enrichment of the heart failure population in atrial fibrillation cases [5]. Indeed, via multi-trait conditional and joint analysis using summary statistics from GWAS of atrial fibrillation, we confirm that the effect of the PITX2 locus on heart failure is explained by its effect in atrial fibrillation (Table 2, Supplemental Table 3). Mendelian randomization (MR) analysis using 110 independent (LD r^2^<0.001) atrial fibrillation-associated variants, provides further evidence for a causal effect of atrial fibrillation on heart failure development (weighted mode MR effect size 0.21 (Odds Ratio 1.23), p<0.0001) (Supplemental Figure 2).

**Table 2.** Lead GWAS variants in multi-trait analysis, Heart Failure sub-phenotypes and echocardiographic traits

| **Sentinel Variant** | **All cause Heart failure**<br>(N=448,549) | **Multi-trait Conditional analysis\***<br>(N=448,549) | **Ischemic Heart failure**<br>(N=27,068) | **Non ischemic Heart failure**<br>(N=27,068) | **HFrEF**<br>(N=18,498) | **HFpEF**<br>(N=18,498) | **LVEF**<br>(N=18,498) | **LVEDD**<br>(N=9,150) | **IVSD**<br>(N=9,577) |
| --- | --- | --- | --- | --- | --- | --- | --- | --- | --- |
| rs580698 | 4.5e-08<br>(0.13) | 6.7e-08 (0.13) | 0.02 (0.15) | 0.005<br>(0.16) | 0.07 (0.18) | 0.81<br>(0.02) | 0.20<br>(-0.10) | 0.05<br>(0.20) | 0.39<br>(0.02) |
| rs1906615 | 2.9e-08<br>(0.10) | 0.005 (0.05) | 0.58<br>(-0.03) | 0.03 (0.10) | 0.08<br>(-0.13) | 0.14<br>(0.09) | 0.69<br>(0.02) | 0.51<br>(-0.05) | 0.23<br>(0.03) |
| rs9411378 | 3.7e-08<br>(0.11) | 1.4e-07 (0.11) | 0.24<br>(0.06) | 0.13<br>(0.08) | 0.36<br>(0.07) | 0.26<br>(0.07) | 0.23<br>(0.08) | 0.10<br>(0.17) | 0.44<br>(-0.02) |

All cell values are formatted as p-value (effect size estimate). N denotes the total sample size for each analysis.
HFpEF: Heart failure with preserved ejection fraction, HFrEF: Heart failure with reduced ejection fraction, IVSD: Interventricular septum diameter, LVEDD: Left ventricular end diastolic diameter, LVEF: Left ventricular ejection fraction.
\*Multi-trait analysis is conditioned on the following Heart failure risk factors: Ischemic Heart Disease, Hypertension, Diabetes Mellitus, Atrial Fibrillation

### Variation in an ACTN2 gene enhancer leads to heart failure and ventricular remodeling

The chromosome 1 locus tagged by the SNP rs580698 is found near the ACTN2 gene, a cardiac sarcolemmal gene at which rare mutations have recently been associated with the development of cardiomyopathy and consequently heart failure [9]. Multi-trait conditional and joint analysis with common heart failure risk factors (atrial fibrillation, ischemic heart disease, hypertension, diabetes mellitus) does not result in a significant change in the effect of the ACTN2 locus on heart failure suggesting that the association signal is not primarily mediated via these other diseases (Table 2, Supplemental Table 3). A phenome-wide association approach (PheWAS) using echocardiographic and other phenotypic information available for a subset of our cohorts and participants demonstrates that the ACTN2 locus is significantly associated with both ischemic and non-ischemic heart failure and has a trend for an effect in left ventricular dilation and heart failure with reduced ejection fraction, thereby suggesting its potential role in mechanisms predisposing to left ventricular adverse remodeling in response to various initial insults (Table 2, Supplemental Figure 3). Chromatin state data for the ACTN2 locus from Roadmap Epigenomics reveal a broad area of muscle specific active enhancer elements in the skeletal muscle, fetal heart, left and right ventricular tissues (Figure 2A, Supplemental Figure 4). Integration with eQTL data does not reveal any compelling evidence of colocalization between the GWAS signal and altered expression of nearby genes in adult blood or post-mortem adult heart tissues (Supplemental Table 4). Conditioning on the sentinel variant eliminates the signal for association of the locus with heart failure, suggesting a single causal variant effect in high LD with the sentinel SNP (Supplemental Figure 5).

**Figure 2.**
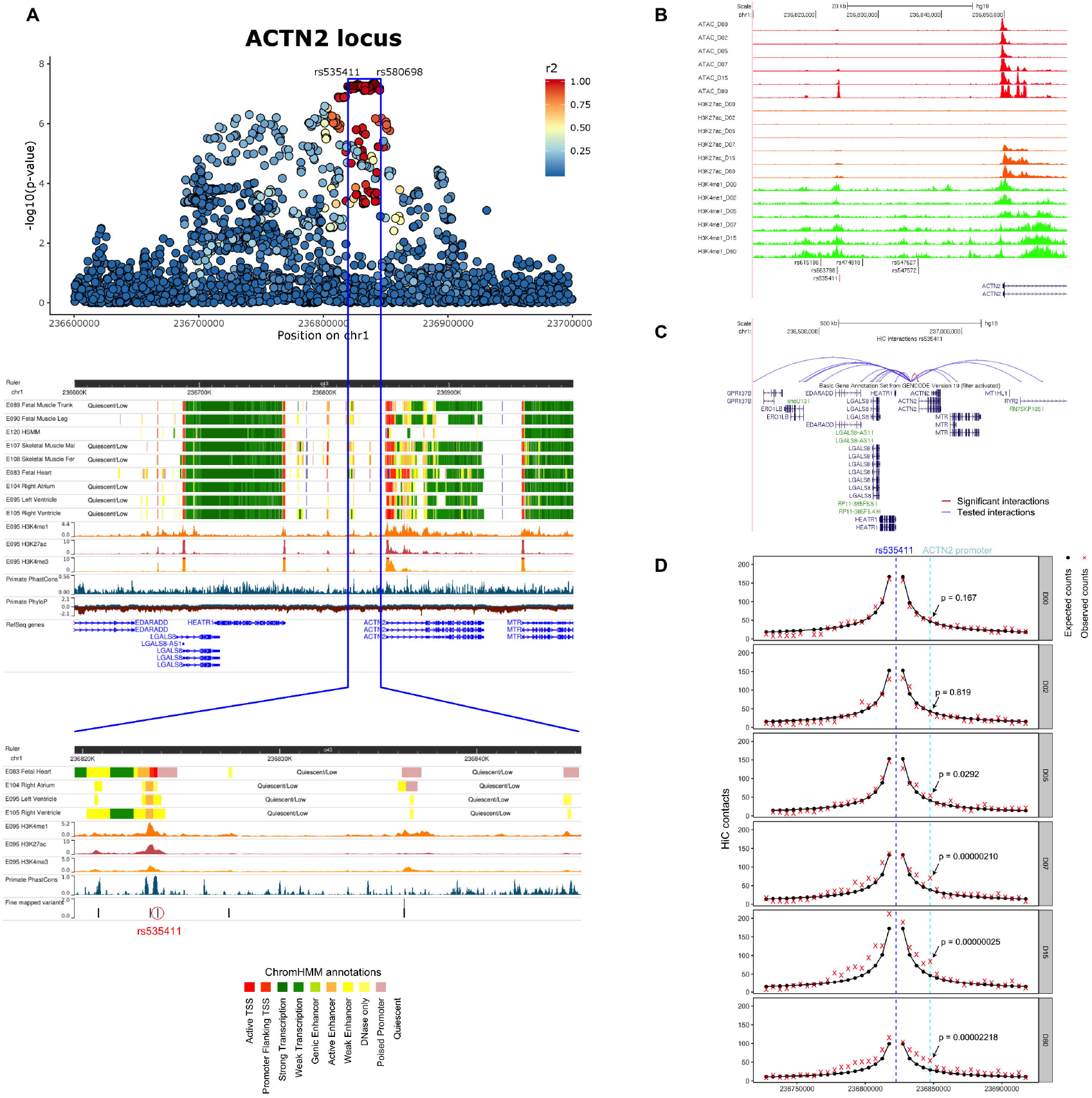
A. Manhattan plot of the ACTN2 locus and the corresponding Roadmap ChromHMM 25-state model annotations in cardiac and muscle cell-types and tissues, H3K4me1, H3K4me3, H3K27ac peaks in left ventricle and PhyloP and PhastCons evolutionary conservation values. The blue box highlights the region that contains all 88 credible set variants identified by CAVIAR and the bottom panel shows a zoomed in view of the credible set region in Roadmap Epigenomics. B. ATAC-seq and ChIP-seq (H3K27ac, H3K4me1) peaks in the hESC-to-cardiomyocyte differentiation model for the fine-mapped SNPs that overlap cardiac specific active chromatin states. We see that one SNP (rs535411) is overlapping an ATAC-seq peak that starts to develop on Day 7 of cardiomyocyte differentiation and remains active until Day 80. Ranges for peak values: ATAC-seq: 0-127, H3K27ac-seq: 0-127, H3K4me1-seq:0-10. C. HiC data from cardiomyocytes on Day 80 of differentiation for the interaction between the 5Kb region containing rs535411 and the promoters of nearby (within 1Mb) genes. Significant interactions after Bonferroni adjustment are colored red, while non-significant interactions (Bonferroni p>0.05) are colored blue. D. HiC data in different stages of cardiomyocyte differentiation centered at rs535411. Black lines/dots: expected interaction counts which follow distance-based decay, Red crosses: observed interaction counts, Blue dashed line: 5Kb region containing rs535411, Cyan dashed line: 5Kb region containing ACTN2 promoter, p-value indicates the upper-tailed Poisson probability with the expected counts as lambda.

Since eQTL analysis in adult tissue did not identify a target gene for the locus, in an effort to provide a credible hypothesis of how this locus is associated with heart failure, we proceeded with additional functional characterization. The first step was to fine map putative causal variants within the locus. First, we generated a credible set of SNPs for the ACTN2 locus based on the GWAS associations and linkage disequilibrium pattern for each region of interest using CAVIAR [10]. Then, we intersected the credible set of SNPs with active chromatin states using the Roadmap Epigenomics ChromHMM 25-state model [11] for cardiac tissues and with candidate cis-regulatory elements (ccREs) from the ENCODE registry [12]. Of 88 variants in almost perfect LD within the credible set for ACTN2, only six overlapped regulatory elements in both Roadmap and ENCODE and were therefore used in downstream analyses (Supplemental Table 5).

Next, we verified the presence of active chromatin states overlapping our ACTN2 locus SNPs in engineered human embryonic stem cell (hESC) derived cardiomyocytes during different stages of their differentiation. We showed that one of the six target variants, rs535411 (Supplemental Table 6) overlaps cardiomyocyte specific ATAC-seq, H3K4me1, and H3K27ac peaks that start to appear on day 7 of hESC differentiation into a cardiomyocyte and persist until at least day 80 (Figure 2B). High resolution chromatin conformation capture (HiC) analysis of our hESC to cardiomyocyte model on day 80 of differentiation shows that the ATAC-seq peak is in contact with the ACTN2 gene promoter (observed/expected interaction frequency = 1.82, p=0.00002) (Figure 2C, Supplemental Table 7) and its interaction is dynamic and increases during differentiation (Figure 2D).

Independent experiments from published literature support the presence of cardiac muscle enhancer in the identified region. Specifically, ChiP-seq data of p300/CREBBP from an independent cardiomyocyte experiment show a peak at the identified region [13] suggestive of chromatin-accessible active regulatory elements. Since the ACTN2 gene is known to be induced during cardiomyocyte maturation [14] and our hESC experiments confirm a dynamic regulatory region that switches on during cardiomyocyte differentiation, the absence of evidence of cis-eQTL effects of our putative causal variant with the ACTN2 gene may reflect a dynamic effect of the enhancer on gene expression during the maturation process. Moreover, prior studies support the role of SNPs in this region in cardiac function. Our fine-mapped variant rs535411 is associated with left ventricular end diastolic dimension (beta= 0.022, p=5.07e-05) in a recent large scale GWAS of echocardiographic traits [15], which corroborates our hypothesis for a role of the locus in left ventricular remodeling. Beyond ACTN2, the data from our genetic correlation analysis (Figure 1B) support a broader role of common variants related to structural musculoskeletal proteins in heart failure by revealing strongly shared heritability between heart failure and multiple musculoskeletal disorders (including osteoarthritis, enthesopathies, intervertebral disk disease) and smooth muscle disorders (esophageal, gastric and duodenal diseases).

### Regulatory variants of the ABO gene predispose to heart failure

Lastly, the chromosome 9 locus tagged by the SNP rs9411378 is found in an intron of the ABO gene, a gene that determines blood type and has been linked to the development of ischemic heart disease [16]. A PheWAS of the sentinel variant across 4,155 GWAS from the GWAS Atlas [17] shows its significant effects in hematologic (red blood cell count, white blood cell count, monocyte cell count, hemoglobin concentration) and metabolic traits (lipid disorders, diabetes, activated partial thromboplastin time) (Supplemental Table 8, Figure 3A), whereas a similar PheWAS approach on 1,448 traits from the UK BioBank reveals its association with venous thromboembolism (Supplemental Figure 6). Interestingly, conditioning on several traits associated with our sentinel variant for which GWAS summary statistics are available or on known heart failure risk factors does not significantly change the signal of association between the ABO locus and heart failure (Table 2, Supplemental Table 3), suggesting a direct effect of the locus on heart failure independent of its effect on other human disorders. The locus did not show any active enhancer or promoter states in cardiac tissues but instead overlapped active enhancer states in primary hematopoietic stem cells and intestinal cells (Supplemental Figure 7 and 8). The sentinel variant was a strong eQTL for ABO gene expression in eQTLGen and GTEx whole blood, consistent with our findings of active chromatin state overlap in hematopoietic lineage cells. In addition, the eQTL signal had strong evidence of colocalization with the GWAS signal for the same locus (posterior probability 96%) (Figure 3B). Notably, the sentinel variant in our GWAS is in LD with the most common variant (rs8176719) associated with O-blood type via a frameshift mutation that is thought to inactivate ABO (LD R^2^ 0.64 in 1000 Genome Europeans). However, the effects of our lead variants on the expression of ABO remain after stratifying by rs8176719 genotype (Figure 3C) suggesting an additional regulatory role for our variants which goes beyond tagging the O-blood type variant rs8176719. Similarly, in our GWAS, a strong signal for association remains within the locus after conditioning on rs8176719 (Supplemental Figure 9). We should note though that rs8176719 is genotyped or accurately imputed only on a small subset of our participants (35,836 individuals), which may limit interpretation of this analysis as definitive evidence of an independent signal. Although having non-O blood type via a structural coding variation in the ABO gene has been linked to cardiovascular disease and cardiovascular mortality [18], the mechanisms underlying this association are not fully understood. Our finding that regulatory variation of the ABO gene’s expression is linked to the development of heart failure highlights the importance of ABO in cardiovascular disease and opens the door to further studies to decipher the cellular mechanisms involved.

**Figure 3.**
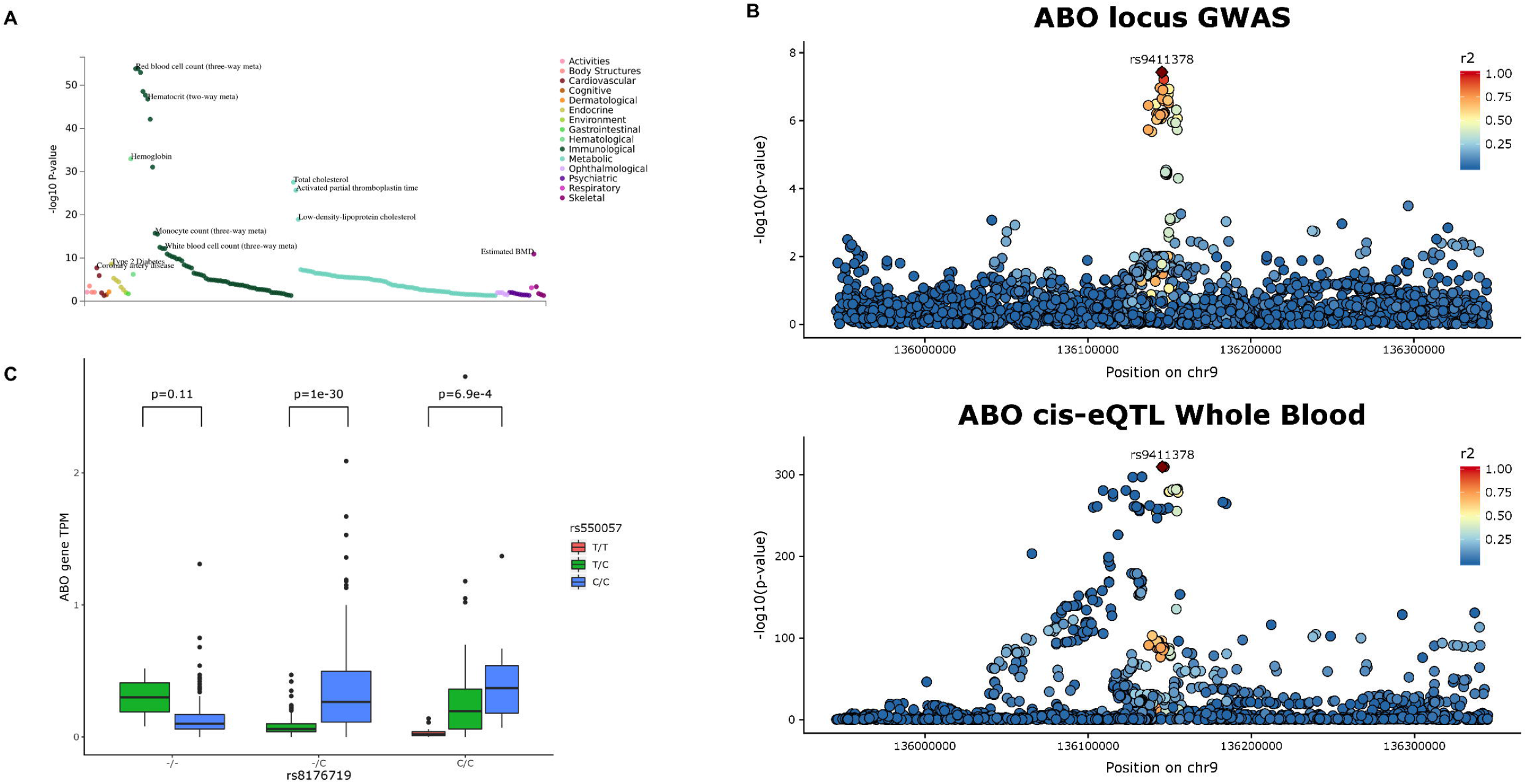
A. Phenome-wide association of the rs9411378 SNP across over 4,000 GWAS from the GWAS Atlas. B. Manhattan plot of the ABO locus overlaying a similar manhattan plot for the ABO gene eQTL in eQTLGen. R^2^ is the 1000 Genomes phase 3 Europeans LD. We see that the signal colocalizes which is confirmed by coloc (posterior probability of colocalization 96%). C. Expression transcripts per million for the ABO gene in GTEx Whole blood stratified by the genotype of rs8176719 (variant defining blood group O) and rs550057 (variant tagging our sentinel variant rs9411378 (LD r^2^=0.92 in 1000 Genomes Europeans)). We see that the T allele for rs550057 is associated with non-O blood group, leads to decreased expression of ABO in individuals that are not blood group O and denotes increased risk of Heart Failure in our GWAS (GWAS beta=0.0961, p-value=6.1e-8).

In summary, we performed the largest genome-wide association study for heart failure to-date and replicated our findings in a similarly powered cohort. Our results validate the use of this approach to discover regulatory variants associated with heart failure predisposition in response to a variety of cardiac insults, reveal a new mechanism for the disease associated with the regulation of a sarcolemmal protein during differentiation, underscore the role of the ABO gene in cardiovascular disease and highlight broadly shared heritability between heart failure and musculoskeletal disorders.

## Methods

### Samples

We performed genome wide association studies in five cohorts that study cardiovascular disease (Framingham Heart Study, Cardiovascular Health Study, Atherosclerosis Risk in Communities Study, Multi-Ethnic Study of Atherosclerosis, Women’s Health Initiative) and the eMERGE initiative. Genotype and phenotype raw data were downloaded from dbGAP (accession numbers phs000007.v29.p11, phs000287.v6.p1, phs000209.v13.p3, phs000280.v4.p1, phs000200.v11.p3, phs000888.v1.p1). For each individual study we performed sample level filtering (excluding samples with assigned and genotype sex discrepancy, extreme deviations from heterozygosity or missingness). We also excluded individuals that were not of European Ancestry and for every group of individuals that were related (Identity by descent (IBD) >0.125) we randomly selected one.

In addition, for each study SNP level filtering was performed to exclude SNPs that had significant deviations from Hardy Weinberg equilibrium in heart failure controls, minor allele frequency less than 0.01, missing call rate > 0.05 and differential missingness between heart failure cases and controls. For studies that analyzed their populations with different genotyping arrays, we also excluded SNPs that had significant deviation in minor allele frequencies (MAF) between the different arrays. For individuals that were genotyped in more than one genotyping array, we selected the array that had the most extensive genotyping. We proceeded with imputing and analyzing each array separately for every study.

### Imputation

We imputed each study to the 1000 Genomes phase 3 reference panel using Minimac3 [19] after pre-phasing with Eagle [20] on the Michigan Imputation Server. Prior to imputation, we lifted all SNPs to the hg19 human genome build using the UCSC liftOver tool, aligned all SNPs to the positive strand and filtered out SNPs whose minor allele frequencies deviated by >0.2 compared to the reference panel’s MAF and SNPs A/T or G/C SNPs with MAF>0.4 as those are prone to strand alignment errors. After imputation, we excluded all imputed SNPs with imputation r squared (INFO score) < 0.7, SNPs with MAF <0.01 and SNPs with Hardy-Weinberg p-value <1e-4. For the eMERGE cohort, imputation was performed independently prior to the start of this study with procedures detailed elsewhere [21] and we subsequently applied the same post-imputation filters.

### Genome wide association

For each study, we performed a GWAS for heart failure controlling for age, sex and the first 10 genotype principal components (PCs). PCs were calculated based on a set of independent (LD r^2^<0.2) genotyped or high-quality imputed SNPs (INFO score>0.9) in an unrelated population (IBD <0.08) and the SNP loadings were subsequently used to calculate the eigenvectors for all individuals included in the analysis. In the eMERGE cohort, since the population was collected from multiple different hospitals across the United States, we included an additional multilevel categorical covariate denoting the sample source. All GWAS were performed using a linear mixed model with the saddlepoint approximation (SAIGE) [22] to account for any residual relatedness structure in our analysis and for case-control imbalance which is inherent in our phenotype of interest. For the UK BioBank cohort we used the summary statistics for all cause Heart Failure (PheCode 428) generated by analyzing the UK BioBank data in the SAIGE paper [22].

### Meta-analysis

We meta analyzed the results of all our GWAS using fixed effects inverse variance meta analysis via the software METAL [23]. We kept only SNPs that were present in at least 3 studies and 5,000 individuals. The following tools were used for the GWAS: Python, R, PLINK [24], SNPRelate [25], SAIGE [22], METAL [23].

### Replication

We replicated our findings in an independent cohort of 24,829 Heart failure cases and 1,614,513 controls of European ancestry within the 23andMe research cohort. Heart failure in the replication population was self-reported as an answer to the question “Have you ever been diagnosed with or treated for Heart failure?”. All three replication variants were imputed with high quality (imputation r^2^>0.95) using an imputation panel that combined the 1000 Genomes Phase 3 panel with the UK10k panel. The variants were analyzed via logistic regression assuming an additive model with covariates for age, sex, the first five genotype PCs, and indicator variables to represent the genotyping platform. The p-values were adjusted for an LD score regression intercept of 1.043.

### Phenome-wide association of heart failure subtypes

For each of the five cohorts in our study and the eMERGE cohort, we classified heart failure individuals as having ischemic heart failure if they also had a history of diagnosed ischemic heart disease, myocardial infarction, percutaneous coronary intervention or coronary artery bypass graft surgery and non-ischemic heart failure otherwise. We also classified individuals as heart failure with reduced ejection fraction if they had heart failure and at least one echocardiogram showing a left ventricular ejection fraction (LVEF) <50%, and heart failure with preserved ejection fraction otherwise. Individuals that did not have information on myocardial infarction history or echocardiographic information were not included in the respective analyses. We also obtained continuous data of LVEF, left ventricular end diastolic diameter and interventricular septum diameter from each individual’s most recent available echocardiogram. Each of our sentinel variants from the general heart failure GWA meta analysis were tested for an effect in each of these variables using SAIGE for the categorical variables and linear regression assuming an additive genotype effect for the continuous variables with the same covariates as in our primary GWAS. The results cross-cohort were meta-analyzed using METAL.

### Other phenotype associations

To evaluate if our lead GWAS variants had associations with other phenotypes we queried the NHGRI-EBI GWAS catalog and also evaluated the GWAS atlas [17] which contains data from 4,155 GWAS across 2,960 unique traits and the 1,488 Electronic Health Record-Derived PheWAS codes from the Michigan Genomics Initiative [22].

### Heritability and Genetic Correlation

We used LD score regression [26] with the 1000 Genomes European reference LD to evaluate the liability scale heritability explained by the common variants in our GWAS assuming a population prevalence of 0.018 [6]. We subsequently analyzed our GWAS together with summary statistics from GWAS studies from the UK biobank [27] using the genetic correlation method of the LD score regression pipeline to quantify the shared heritability between our phenotype and other traits [28]. For the genetic correlation analysis we selected traits to analyze based on the following procedure:

1. Among all summary statistics analyzed in the SAIGE paper [22], we first excluded the categories Injuries and poisonings (as it is unlikely to have a major heritable component) as well as Symptoms and Pregnancy complications (as they are too general to have a meaningful interpretation of genetic correlation).
2. We excluded general disease bundles that include the work “other” or “NOS” (eg. Other infectious and parasitic diseases) or are a sign/symptom (eg. Hematuria) or medication (eg. Chemotherapy).
3. We reclassified all infections into the “Infectious Diseases” category and all congenital anomalies to their respective organ system.
4. From every organ system or general disease category we selected the three diseases with the highest number of cases.
5. For every selected disease, we excluded diseases and disorders that are subsets of the same disease or highly related (eg. Selected disease: Hypertension - excluded disease: Essential Hypertension).
6. We excluded diseases whose z-score of observed heritability calculated via LD score regression was <1.

### Conditional analysis based on summary statistics

We used the COJO package [29] from the GCTA pipeline to evaluate the residual association signal within our genome-wide significant loci after conditioning on our sentinel variants or other variants of interest using as reference the LD of the eMERGE heart failure dataset.

### Multi-trait conditional and joint analysis

We used the mtCOJO package [30] from the GCTA pipeline to evaluate the effects of our variants conditioned to other heart failure risk factors (eg. hypertension, atrial fibrillation, ischemic heart disease) and conditions associated with our sentinel variants in PheWAS studies using the 1000 Genomes Europeans reference LD scores.

### Mendelian Randomization analysis

We used the MR base package [31] to perform Mendelian Randomization analysis in order to evaluate the effect of atrial fibrillation on the development of heart failure using summary statistics from a large-scale GWAS meta-analysis of atrial fibrillation [32]. The polygenic risk score was constructed using independent variants (LD r^2^ < 0.001) at a genome-wide significance threshold (p < 5e-8) using as reference the LD of the 1000 Genomes European samples.

### Variant fine-mapping

We followed a step-wise approach based on epigenomic annotations and LD structure for our variant fine-mapping efforts. We first used the Roadmap epigenomics ChromHMM 25-state model [11] across all tested cell types and tissues to visualize our significant loci and identify broad patterns of active promoter or enhancer elements across tissues. We subsequently used CAVIAR [10] with the 1000 Genomes European reference LD and the assumption of at most two causal variants per locus to generate a credible set of SNPs for each locus. The CAVIAR analysis for each locus included all SNPs within 100 kilobases of the locus sentinel SNP. Then, we intersected the SNPs in each credible set with active enhancer or promoter elements predicted by Roadmap epigenomics for heart tissues (fetal heart, left ventricle, right ventricle and right atrium). Finally, we intersected SNPs selected by the previous step with candidate cis-regulatory elements predicted by ENCODE [12]. Bedtools [33] was used for all intersection tests.

### Cardiomyocyte differentiation model

To further probe the effects of the ACTN2 locus on cardiomyocyte function, we used an engineered human embryonic stem cell differentiation model. The assays are fully described elsewhere [34], but in brief, we performed ATAC-seq, ChIP-seq, RNA-seq and HiC experiments in an engineered H9 hESC MLC2v:H2B-GFP reporter transgenic line that was differentiated into cardiomyocytes using a well-stablished Wnt-based cardiomyocyte differentiation protocol [35]. Cardiac cell populations were collected and analyzed at different differentiation stages (Day 0, 2,5,7,15 and 80). We queried our fine-mapped variants by intersecting them with ATAC-seq, H3K4me3 and H3K27ac peaks in the cardiomyocyte differentiation model and we subsequently assessed the HiC contacts between the identified peaks and nearby genes. HiC contacts were generated at 5kb resolution. Expected contacts for each bin are calculated as the genome-wide average of contacts of the same distance, as Hi-C contacts follow a distance-based delay. The observed/expected value for each bin shows the enrichment of HiC contacts relative to the background. We tested the significance of enrichment of observed contacts with respect to the expected contacts using an upper-tail Poisson test with x equals observed contacts and lambda equals expected contacts [36].

### Expression quantitative trait loci (eQTL)

We assessed the effect of our genome wide significant variants in gene expression of nearby genes using two databases: 1) Genotype Tissue Expression (GTEx): We obtained whole genome sequencing and RNA sequencing data from GTEx version 8. We followed the standard pipeline proposed by GTEx v7 [37] to normalize gene expression and perform cis-eQTL analyses. In brief, we filtered out genes with <6 reads or <0.1 counts per million (cpm) in >20% of participants per tissue, performed normalization of expression values between samples using TMM [38] and for each gene, we normalized gene expression across samples by an inverse rank-based transform to the standard normal distribution. The effect of a variant in gene expression was analyzed using linear regression as implemented in MatrixEQTL [39] using age, sex, RNA-seq platform, five genotype PCs and 60 Probabilistic Estimation of Expression Residuals (PEER) factors [40] as covariates. Using the methods above, we tested the effect of the ACTN2 locus fine-mapped variants to the expression of genes within 1 megabase in Left Ventricle tissue and the effect of the ABO locus in Whole blood. 2) To increase the power of detecting a cis-eQTL association in Whole Blood, we obtained cis-eQTL summary statistics from eQTLGen which includes eQTL data from 31,684 samples [41]. We queried our ABO locus sentinel variant in the dataset.

### Colocalization analysis

For each identified significant eQTL result for our variants, we evaluated whether the eQTL and GWAS signals colocalize using a Bayesian colocalization method as implemented in coloc [42] to estimate the posterior probability of an identical causal variant per locus between eQTL and GWAS. Colocalization Manhattan plots for Supplemental Figure 1 were generated using LocusZoom [43].

## Data Availability

GWAS summary statistics will be available upon manuscript publication in a peer reviewed journal

## Acknowledgements

We would like to thank the research participants and employees of 23andMe for making this work possible. The following members of the 23andMe Research Team contributed to this study: Michelle Agee, Stella Aslibekyan, Robert K. Bell, Katarzyna Bryc, Sarah K. Clark, Sarah L. Elson, Kipper Fletez-Brant, Pierre Fontanillas, Nicholas A. Furlotte, Pooja M. Gandhi, Karl Heilbron, Barry Hicks, David A. Hinds, Karen E. Huber, Ethan M. Jewett, Yunxuan Jiang, Aaron Kleinman, Keng-Han Lin, Nadia K. Litterman, Jennifer C. McCreight, Matthew H. McIntyre, Kimberly F. McManus, Joanna L. Mountain, Sahar V. Mozaffari, Priyanka Nandakumar, Elizabeth S. Noblin, Carrie A.M. Northover, Jared O’Connell, Steven J. Pitts, G. David Poznik, J. Fah Sathirapongsasuti, Anjali J. Shastri, Janie F. Shelton, Suyash Shringarpure, Chao Tian, Joyce Y. Tung, Robert J. Tunney, Vladimir Vacic, Xin Wang, Amir S. Zare.

Dr. Arvanitis was supported by NIH T32-HL007227 for this work.

## Author Contributions

Dr. Arvanitis conceptualized and designed the study, performed the analyses, interpreted the results and wrote the manuscript.

Drs. Zhang, Chi and Ren performed the experiments and analyses of the HiC cardiomyocyte differentiation data, interpreted the results and reviewed and revised the manuscript.

Drs. Wang, Auton and the 23andMe Research Team performed the analyses pertaining to GWAS replication and reviewed and revised the manuscript.

Dr. Keramati contributed to the design of the primary GWAS meta-analysis and reviewed and revised the manuscript.

Dr. Post contributed to the design of the study, the interpretation of the results and reviewed and revised the manuscript.

Dr. Battle conceptualized and designed the study, provided oversight for the analyses, interpreted the results and reviewed and revised the manuscript.

## Notes

### Competing Interest Statement

The authors have declared no competing interest.

### Author Declarations

All relevant ethical guidelines have been followed and any necessary IRB and/or ethics committee approvals have been obtained.

Any clinical trials involved have been registered with an ICMJE-approved registry such as ClinicalTrials.gov and the trial ID is included in the manuscript.

